# Digital ethnicity data in population-wide electronic health records in England: a description of completeness, coverage, and granularity of diversity

**DOI:** 10.1101/2022.11.11.22282217

**Authors:** Marta Pineda-Moncusí, Freya Allery, Antonella Delmestri, Thomas Bolton, John Nolan, Johan Thygesen, Alex Handy, Amitava Banerjee, Spiros Denaxas, Christopher Tomlinson, Alastair K Denniston, Cathie Sudlow, Ashley Akbari, Angela Wood, Gary S Collins, Irene Petersen, Kamlesh Khunti, Daniel Prieto-Alhambra, Sara Khalid, the CVD-COVID-UK/COVID-IMPACT Consortium

## Abstract

**Background:** The link between ethnicity and healthcare inequity, and the urgency for better data is well-recognised. This study describes ethnicity data in nation-wide electronic health records in England, UK.

**Methods:** We conducted a retrospective cohort study using de-identified person-level records for the England population available in the National Health Service (NHS) Digital trusted research environment. Primary care records (GDPPR) were linked to hospital and national mortality records. We assessed completeness, consistency, and granularity of ethnicity records using all available SNOMED-CT concepts for ethnicity and NHS ethnicity categories.

**Findings:** From 61.8 million individuals registered with a primary care practice in England, 51.5 (83.3%) had at least one ethnicity record in GDPPR, increasing to 93·9% when linked with hospital records. Approximately 12·0% had at least two conflicting ethnicity codes in primary care records. Women were more likely to have ethnicity recorded than men. Ethnicity was missing most frequently in individuals from 18 to 39 years old and in the southern regions of England. Individuals with an ethnicity record had more comorbidities recorded than those without. Of 489 SNOMED-CT ethnicity concepts available, 255 were used in primary care records. Discrepancies between SNOMED-CT and NHS ethnicity categories were observed, specifically within “Other-” ethnicity groups.

**Interpretation:** More than 250 ethnicity sub-groups may be found in health records for the English population, although commonly categorised into “White”, “Black”, “Asian”, “Mixed”, and “Other”. One in ten individuals do not have ethnicity information recorded in primary care or hospital records. SNOMED-CT codes represent more diversity in ethnicity groups than the NHS ethnicity classification. Improved recording of self-reported ethnicity at first point-of-care and consistency in ethnicity classification across healthcare settings can potentially improve the accuracy of ethnicity in research and ultimately care for all ethnicities.

**Funding:** British Heart Foundation Data Science Centre led by Health Data Research UK.

**Research in context:** *Evidence before this study:* Ethnicity has been highlighted as a significant factor in the disproportionate impact of SARS-CoV-2 infection and mortality. Better knowledge of ethnicity data recorded in real clinical practice is required to improve health research and ultimately healthcare. We searched PubMed from database inception to 14^th^ July 2022 for publications using the search terms “ethnicity” and “electronic health records” or “EHR,” without language restrictions. 228 publications in 2019, before the COVID-19 pandemic, and 304 publications between 2020 and 2022 were identified. However, none of these publications used or reported any of over 400 available SNOMED-CT concepts for ethnicity to account for more granularity and diversity than captured by traditional high-level classification limited to 5 to 9 ethnicity groups.

*Added value of this study:* We provide a comprehensive study of the largest collection of ethnicity records from a national-level electronic health records trusted research environment, exploring completeness, consistency, and granularity. This work can serve as a data resource profile of ethnicity from routinely-collected EHR in England.

*Implications of all the available evidence:* To achieve equity in healthcare, we need to understand the differences between individuals, as well as the influence of ethnicity both on health status and on health interventions, including variation in the behaviour of tests and therapies. Thus, there is a need for measurements, thresholds, and risk estimates to be tailored to different ethnic groups. This study presents the different medical concepts describing ethnicity in routinely collected data that are readily available to researchers and highlights key elements for improving their accuracy in research. We aim to encourage researchers to use more granular ethnicity than the than typical approaches which aggregate ethnicity into a limited number of categories, failing to reflect the diversity of underlying populations. Accurate ethnicity data will lead to a better understanding of individual diversity, which will help to address disparities and influence policy recommendations that can translate into better, fairer health for all.

## Introduction

Health inequity is described by disparities in health status between individuals, such as prevalence of comorbidities, life expectancy, access to and quality of care services and treatments, and risk behaviours such as smoking and alcohol consumption. These factors can be influenced by age, sex, ethnicity, disability, socio-economic status, geographical location, and education, among others.^1^ For example, many of these determinants were risk factors for infection severity, complications, and mortality during the COVID-19 pandemic.^2,3^

“Ethnicity” commonly refers to terms used to self-report an individual’s own perceived ethnic group and cultural background. This multidimensional, evolving concept can comprise physical appearance, culture, language, religion, nationality and identity elements. Although pre- and post-pandemic inequalities by ethnicity are well-described, it is difficult to measure the impact of ethnicity due to poor and variable coding of ethnicity across studies, aggregation of estimates due to low sample sizes and unclear generalisability of research cohorts.

Research and development in healthcare often relies on the quality of data recorded by healthcare professionals in primary (e.g., GP practices) and secondary (e.g., hospitals) care settings. Self-reported variables, such as smoking or ethnicity, are not always collected as healthcare professionals may not request or record them, and individuals can decline to provide this information. When recorded, ethnicity is often inaccurately coded.^4-6^ For instance, a prior study estimated that 41.2% of ethnicity records from non-White British individuals in England hospitals were incorrect.^5^ Ethnicity classifications also change over time, limiting comparability with population-level census data.^7^ Although there are hundreds of heterogenous sources of ethnicity concepts (e.g., SNOMED-CT codes, Read codes), they are commonly collapsed into five or six categories, in part due to power considerations where fine-grained or granular categories would have smaller sample sizes.^8-11^ Nonetheless this oversimplification can result in loss of diversity and precision in studies using ethnicity. Incorrect or unrepresentative ethnicity records risk introducing bias in insights drawn from the literature, leading to inappropriate healthcare. Use of population-wide routinely-collected data offers an opportunity to study non-majority ethnicity groups with sufficient power and at a higher resolution, enabling more diverse ethnicities to be included in research.^10-12^

Inequality between different ethnicities has been highlighted as a significant issue as people from ethnically diverse backgrounds were disproportionately affected by SARS-CoV-2.^11^ To improve the understanding of how ethnicity is recorded, mapped, and used in England, we explored ethnicity records for completeness, consistency, and granularity using data made available via the CVD-COVID-UK/COVID-IMPACT Consortium’s instance of the National Health Service (NHS) Digital Trusted Research Environment (TRE) for England.

## Methods

### Data sources and linkages

This study focused on the General Practice Extraction Service (GPES) Data for Pandemic Planning and Research (GDPPR) data sources,^13,14^ accessible through the NHS Digital TRE, ^15,16^ and uses the linkage to Hospital Episode Statistics (HES) and the UK Office for National Statistics (ONS). Further details are displayed in *Supplemental data*.

### Data access

A data sharing agreement issued by NHS Digital for the CVD-COVID-UK/COVID-IMPACT research programme (ref: DARS-NIC-381078-Y9C5K) enables accredited, approved researchers from institutions party to the agreement to access data held within the NHS Digital TRE service for England.

### Study settings and participants

This retrospective cohort study included all individuals with a unique patient pseudoidentifier in GDPPR from 1^st^ Jan 1997 until 23^rd^ April 2022. Individuals with an invalid age (i.e., age <0 or ≥115 years old) or missing sex were excluded.

### Source codes for ethnicity

Ethnicity is recorded using the following medical concepts (Figure 1):

**Figure 1.**
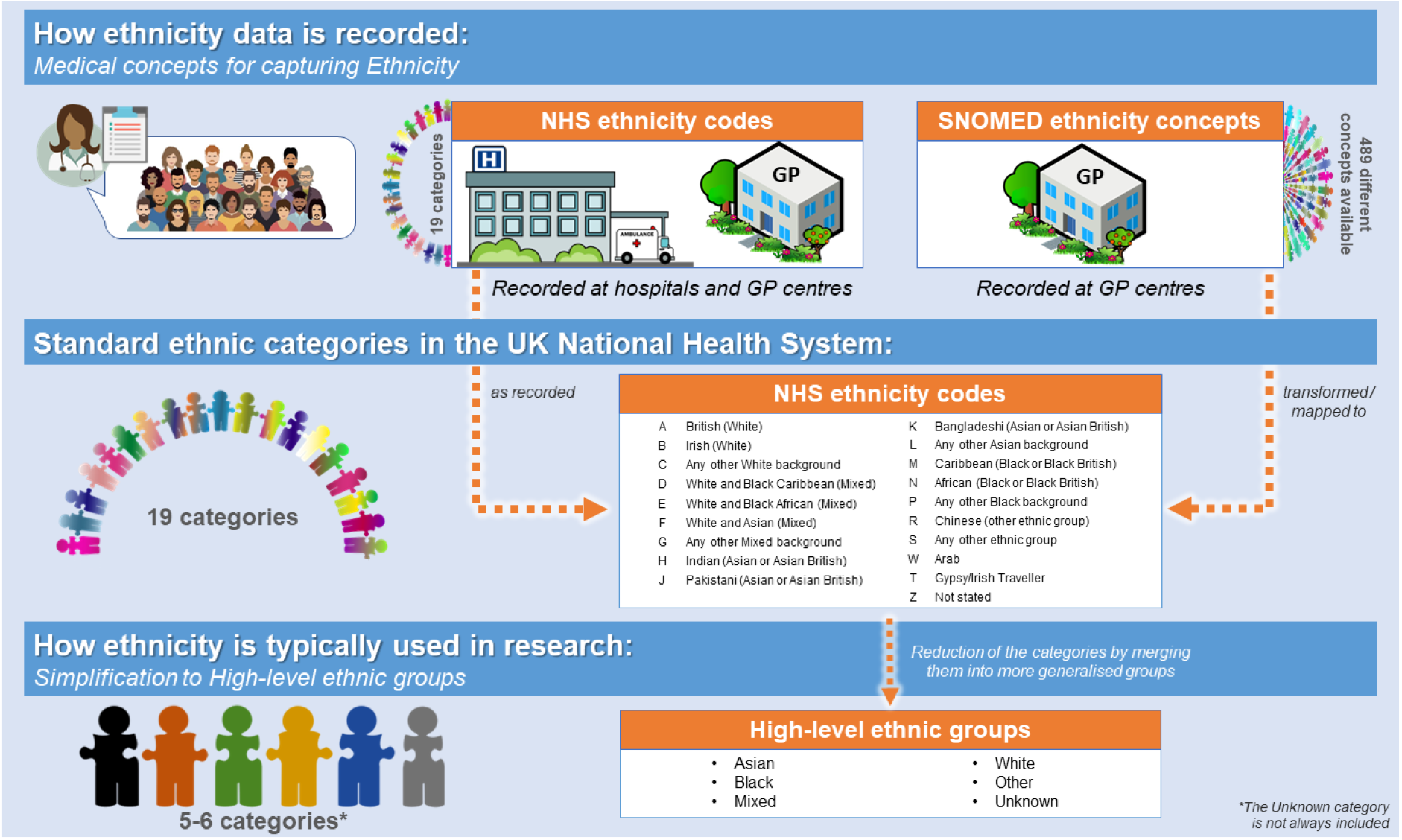
How ethnicity is collected in the UK and typically used for research. Abbreviations: Abbreviations: High-level ethnic groups, general ethnicity classification groups from the Office for National Statistics commonly used in research; NHS, National Health Service in the UK; SNOMED, SNOMED-CT records containing ethnicity concepts.

SNOMED concepts: Standardised vocabulary for digital recording patients’ clinical information across NHS practices and healthcare providers, including ethnicity coding.^17^ We focused on GDPPR records containing Systematized Nomenclature of Medicine Clinical Terms (SNOMED-CT)^18^ UK Edition ethnicity concepts. Any mention of SNOMED concepts in this paper directly refers to these codes.

NHS ethnicity codes: Standard ethnicity categories defined in the NHS Digital Data Dictionary.^19^ Ethnic fields in the NHS tables may use different census classifications, and thus, NHS ethnicity code notation can slightly differ. Table 1 summarises the NHS ethnicity codes available in GDPPR and the corresponding categories in HES and different-year censuses from the UK ONS. Mapping from SNOMED concepts to NHS ethnicity codes was provided by NHS Digital (*Appendix SNOMED mapping*).

**Table 1.**
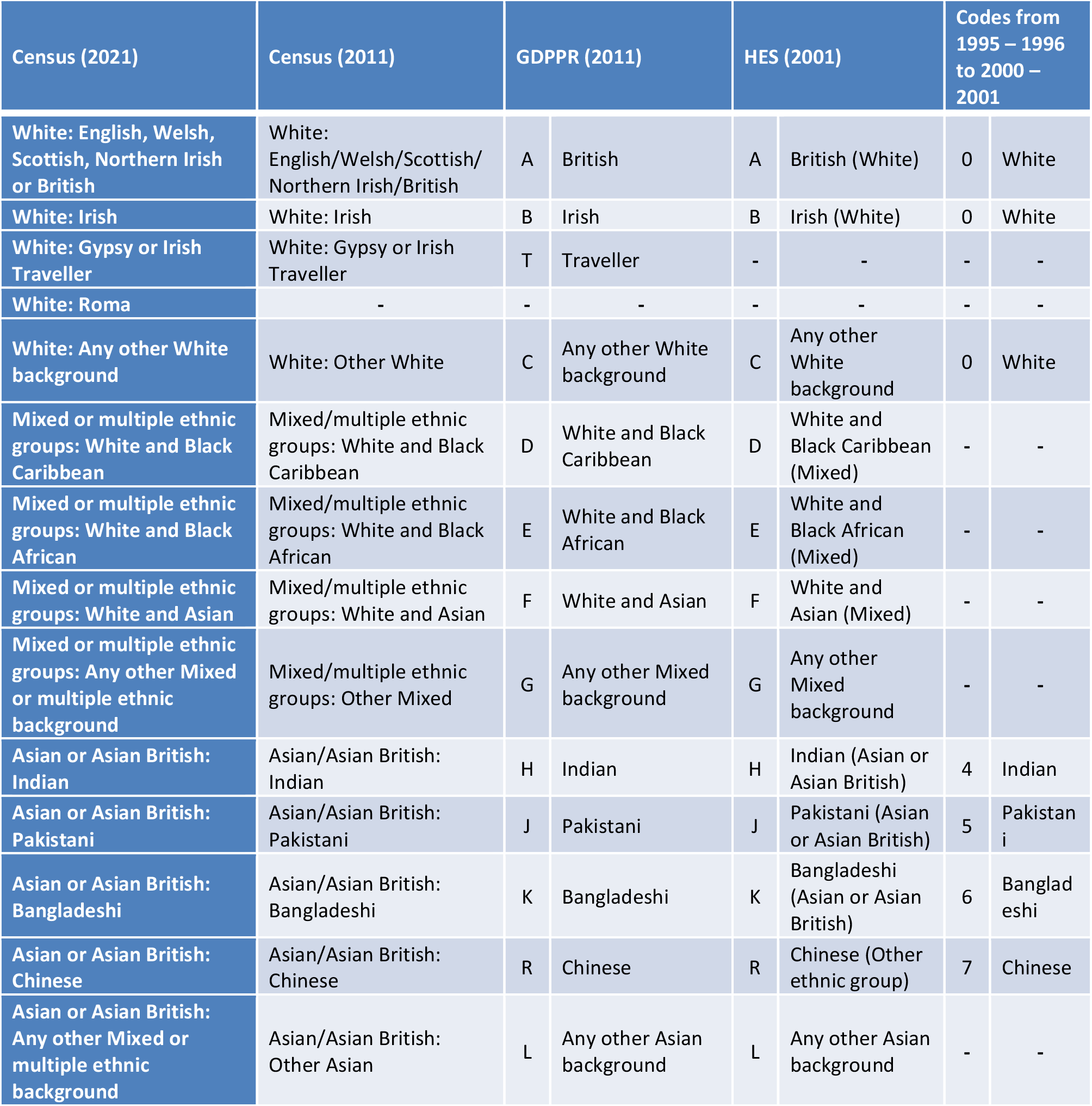
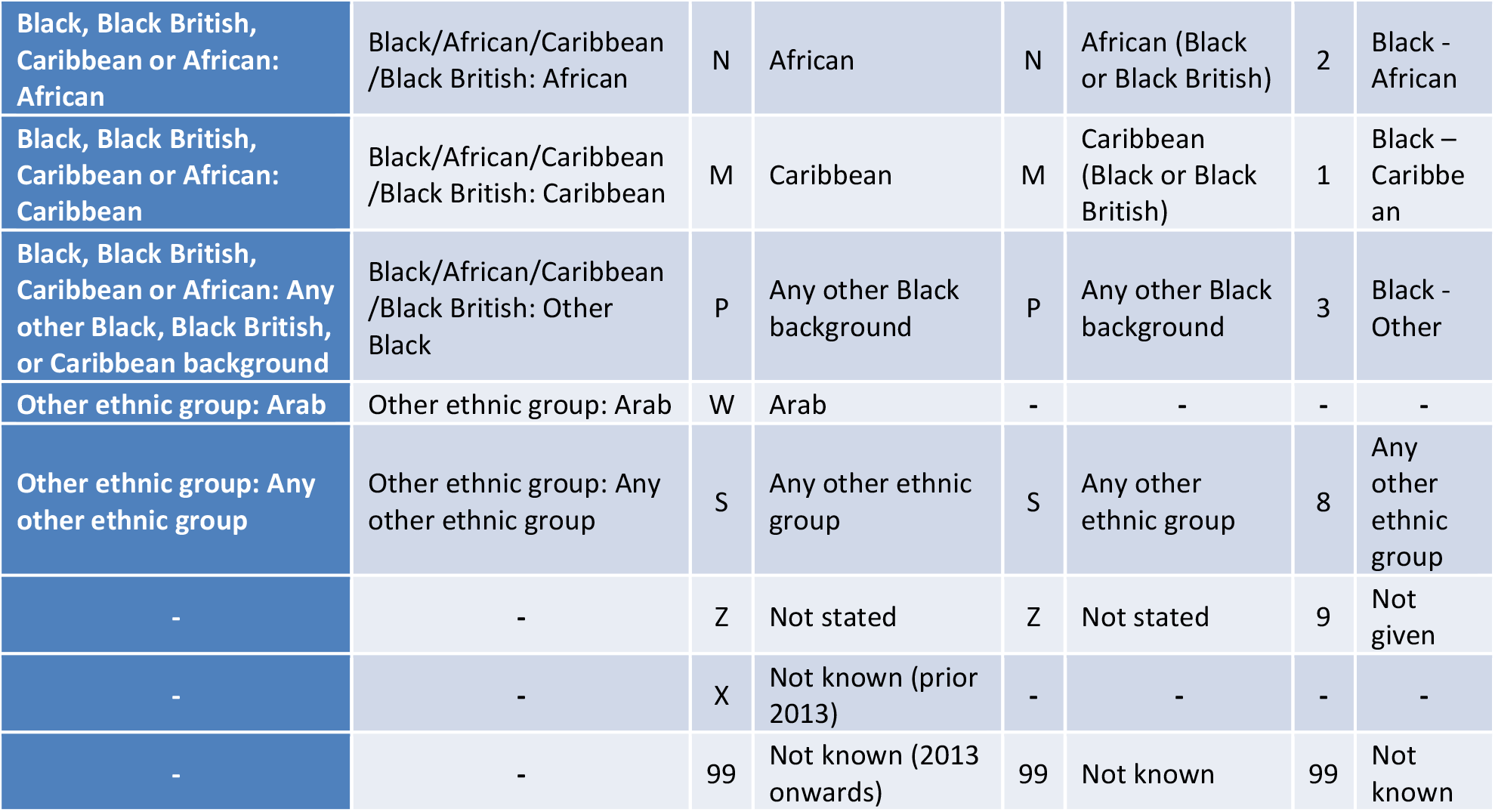
NHS ethnicity codes for Ethnicity (A-Z) available in GDPPR and corresponding categories in Census (2021), Census (2011), Hospital Episode Statistics, and notation before 2001. The Hospital Episode Statistics (HES) data uses census notation based on the 2001 Census. ‘Z: not stated’ indicates that the person was asked and either refused to provide this information or was genuinely unable to choose a response. ‘X: Not known’ indicates that the person was not asked or was not in a condition to be asked (e.g., unconscious). Abbreviations: NHS, National Health Statistics.

An individual’s ethnicity may be recorded using either SNOMED-CT concepts or NHS ethnicity codes in GDPPR (primary care records), whereas it may only be recorded using the latter in NHS HES admitted patient records (i.e., hospital records)

### Other ethnicity classifications

High-level ethnic groups: Asian/Asian British, Black/African/Caribbean/Black British, Mixed, Other Ethnic Groups, Unknown, and White. Based on ONS ethnic group high-level category descriptions.^20,21^

The algorithm used within the TRE to condense NHS ethnicity codes and SNOMED concepts to these classifications is provided in *Supplemental data*.

### Covariates

Death date was obtained through civil registration death table which is curated by the ONS and records primary and secondary causes of death using ICD-10. All additional characteristics of individuals were extracted from GDPPR data. Full description of additional characteristics is available in *Supplemental data*.

### Patient and public involvement

The UK National Institute for Health Research-British Heart Foundation (BHF) Cardiovascular Partnership lay panel comprising individuals affected by cardiovascular disease reviewed and approved this project.

### Statistical analysis

#### Completeness: missing ethnicity data

To study completeness, individual-level ethnicity data were extracted from GDPPR using SNOMED concepts and/or NHS ethnicity codes, prioritising SNOMED concepts when available. For individuals with missing ethnicity data in GDPPR, we extracted HES-linked ethnicity data using NHS ethnicity codes (Supplemental Figure 1).

Ethnicity missingness was defined as (i) no record in either GDPPR or HES or (ii) an ethnicity code of “not stated” referring to individuals who were asked but preferred not to state their ethnicity and individuals who did not know what to answer. We compared the clinical characteristics of GDPPR individuals whose ethnicity data were obtained from GDPPR, were linked from HES, or were not recorded.

#### Inconsistency: multiple records

To study the presence of multiple ethnicity codes co-existing in the same patient in GDPPR records, individual’s SNOMED concepts were converted into the corresponding 19 NHS ethnicity code categories. Each pair of NHS ethnicity code ethnicities were summed to determine their frequency in individuals with multiple ethnicity codes. If a patient had more than two codes assigned, a count of one was added for each pairing. We compared the prevalence of individuals with multiple codes in GDPPR and HES.

#### Inconsistency: potential discrepancies between classifications

To study potential discrepancies and misclassifications between the different ethnicity classifications, we explored mappings between SNOMED and NHS ethnicity codes and between NHS ethnicity codes and high-level ethnic groups.

#### Granularity: from high-level categories to SNOMED concepts

Here “granularity” refers to the degree of detail, i.e., sub-groups within an ethnicity group. Definitions of the most recent SNOMED record in GDPPR individuals were explored.

Data were prepared using Python V.3.7 and Spark SQL (V.2.4.5) on Databricks Runtime V.6.4 for Machine Learning. Data were analysed using Python in Databricks and RStudio (Professional) Version 1.3.1093.1 driven by R Version 4.0.3. All code for data preparation and analysis are available on GitHub (https://github.com/BHFDSC/CCU037_01).

## Results

### Completeness of ethnicity data

We identified 61,810,570 individuals with unique identifiers in the GDPPR dataset on 23^rd^ April 2022. We excluded 403 of these individuals for invalid age or missing sex. Ethnicity records were present for 51,493,087 (83·3%) of the remaining individuals. This 16·7% of missing ethnicity data in GDPPR was reduced to 6·1% (n=3,757,574) when supplemented with ethnicity data from linked HES (Figure 2).

**Figure 2.**
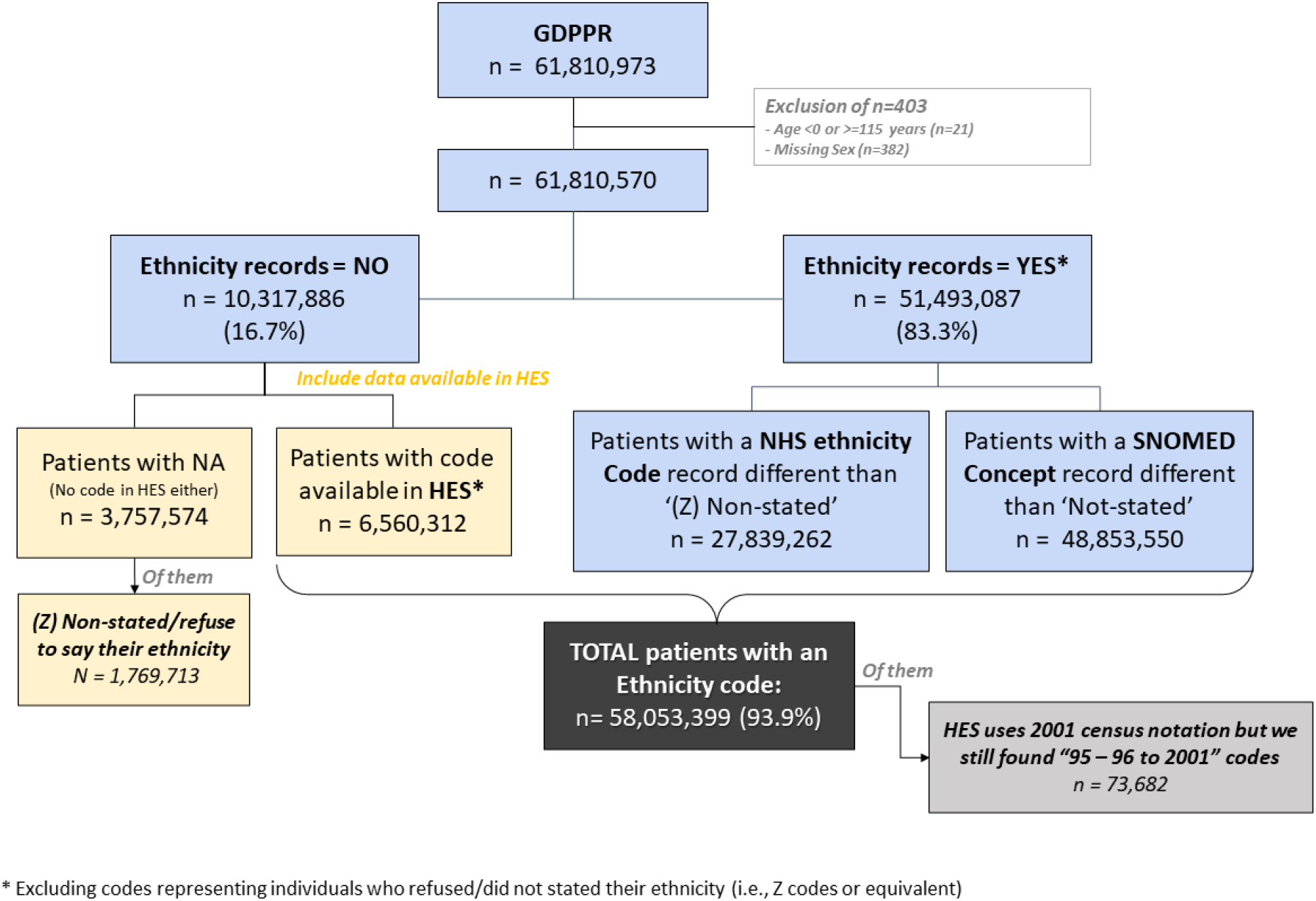
Flow chart of availability of ethnicity records for individuals present in GDPPR. Abbreviations: GDPPR, General Practice Extraction Service (GPES) Data for Pandemic Planning and Research; HES, hospital episode statistics; NHS, National Health Service in the UK; SNOMED, SNOMED-CT records containing ethnicity concepts; NA, not available ethnicity.

Individuals with missing ethnicity data were generally younger, with a median age [Q1, Q3] of 35·0 [22·0, 53·0] years vs 42·0 [24·0, 61·0] years for those with ethnicity from GDPPR and 36·0 [18·0, 58·0] years for those with ethnicity linked from HES. A greater proportion of those with missing ethnicity were male (58·6%) than those with ethnicity from GDPPR (48·9% male) or HES (54·0% male). They also had fewer comorbidities (Table 2) and a greater proportion came from the South East and South West regions of England (Supplemental Figure 2). Individuals aged 18-29 years had between 5·7 and 9·0 percentage points more missing ethnicity data than any other age group (Table 2).

**Table 2.** Comparison of individuals with and without an ethnicity record in GDPPR or linked from HES.

| Individuals with Characteristics | Ethnicity recorded in GDPPR* (N=51,493,087) | Ethnicity linked from HES* (N=6,560,312) | Ethnicity not stated/recorded** (N=3,757,171) | Total GDPPR (N=61,810,570) |
| --- | --- | --- | --- | --- |
| **Age, years:** |  |  |  |  |
| Mean (SD) | 42·6 (23·5) | 39·1 (25·1) | 38·1 (20·9) | 41·9 (23·6) |
| Median [Min, Max] | 42·0 [0, 115] | 36·0 [0, 113] | 35·0 [0, 115] | 41·0 [0, 115] |
| **Age groups, years:** |  |  |  |  |
| 0-17 | 9345962 (18·2%) | 1539936 (23·5%) | 634290 (16·9%) | 11520188 (18·6%) |
| 18-29 | 7203161 (14·0%) | 1137551 (17·3%) | 866021 (23·0%) | 9206733 (14·9%) |
| 30-39 | 7622783 (14·8%) | 859347 (13·1%) | 645268 (17·2%) | 9127398 (14·8%) |
| 40-49 | 6809996 (13·2%) | 707735 (10·8%) | 496539 (13·2%) | 8014270 (13·0%) |
| 50-59 | 6966406 (13·5%) | 777275 (11·8%) | 461384 (12·3%) | 8205065 (13·3%) |
| 60-69 | 5706615 (11·1%) | 601874 (9·2%) | 330270 (8·8%) | 6638759 (10·7%) |
| 70-79 | 4613388 (9·0%) | 468309 (7·1%) | 195175 (5·2%) | 5276872 (8·5%) |
| 80-89 | 2462254 (4·8%) | 328554 (5·0%) | 94656 (2·5%) | 2885464 (4·7%) |
| 90+ | 762522 (1·5%) | 139731 (2·1%) | 33568 (0·9%) | 935821 (1·5%) |
| **Sex:** |  |  |  |  |
| Female | 26311290 (51·1%) | 3015261 (46·0%) | 1554863 (41·4%) | 30881414 (50·0%) |
| Male | 25181797 (48·9%) | 3545051 (54·0%) | 2202308 (58·6%) | 30929156 (50·0%) |
| **BMI (kg/m2):** |  |  |  |  |
| Mean (SD) | 27·3 (6·63) | 27·3 (6·80) | 26·4 (6·67) | 27·2 (6·65) |
| Median [Min, Max] | 26·4 [10·0, 80·0] | 26·5 [10·0, 80·0] | 25·5 [10·0, 79·9] | 26·4 [10·0, 80·0] |
| Missing | 26413905 (51·3%) | 4731250 (72·1%) | 2905792 (77·3%) | 34050947 (55·1%) |
| **IMD decile:** |  |  |  |  |
| 1 (most deprived) | 5030900 (9·8%) | 611062 (9·3%) | 290292 (7·7%) | 5932254 (9·6%) |
| 2 | 5002481 (9·7%) | 585319 (8·9%) | 310009 (8·3%) | 5897809 (9·5%) |
| 3 | 5026837 (9·8%) | 586855 (8·9%) | 327026 (8·7%) | 5940718 (9·6%) |
| 4 | 4821880 (9·4%) | 630567 (9·6%) | 345865 (9·2%) | 5798312 (9·4%) |
| 5 | 4670559 (9·1%) | 623723 (9·5%) | 331139 (8·8%) | 5625421 (9·1%) |
| 6 | 4657873 (9·0%) | 627814 (9·6%) | 354218 (9·4%) | 5639905 (9·1%) |
| 7 | 4489880 (8·7%) | 620733 (9·5%) | 340289 (9·1%) | 5450902 (8·8%) |
| 8 | 4467923 (8·7%) | 632435 (9·6%) | 338721 (9·0%) | 5439079 (8·8%) |
| 9 | 4365799 (8·5%) | 634458 (9·7%) | 338340 (9·0%) | 5338597 (8·6%) |
| 10 (less deprived) | 4292948 (8·3%) | 640425 (9·8%) | 362403 (9·6%) | 5295776 (8·6%) |
| Missing | 4666007 (9·1%) | 366921 (5·6%) | 418869 (11·1%) | 5451797 (8·8%) |
| **Geographic regions\*\*\*:** |  |  |  |  |
| East Midlands | 3471792 (6·7%) | 336184 (5·1%) | 234275 (6·2%) | 4042251 (6·5%) |
| East of England | 4462265 (8·7%) | 480993 (7·3%) | 373491 (9·9%) | 5316749 (8·6%) |
| London | 8133607 (15·8%) | 743482 (11·3%) | 541694 (14·4%) | 9418783 (15·2%) |
| North East | 1841607 (3·6%) | 211971 (3·2%) | 95445 (2·5%) | 2149023 (3·5%) |
| North West | 5975649 (11·6%) | 1024996 (15·6%) | 336139 (8·9%) | 7336784 (11·9%) |
| South East | 6678501 (13·0%) | 1163911 (17·7%) | 645049 (17·2%) | 8487461 (13·7%) |
| South West | 3484388 (6·8%) | 741784 (11·3%) | 336138 (8·9%) | 4562310 (7·4%) |
| West Midlands | 4994436 (9·7%) | 643818 (9·8%) | 235647 (6·3%) | 5873901 (9·5%) |
| Yorkshire and The Humber | 4773116 (9·3%) | 460537 (7·0%) | 319327 (8·5%) | 5552980 (9·0%) |
| Missing | 7677726 (14·9%) | 752636 (11·5%) | 639966 (17·0%) | 9070328 (14·7%) |
| **Death** | 995430 (1·9%) | 238372 (3·6%) | 39434 (1·0%) | 1273236 (2·1%) |
| **Smoke use record** | 20509149 (39·8%) | 2305227 (35·1%) | 1064288 (28·3%) | 23878664 (38·6%) |
| **Heavy alcohol use** | 796064 (1·5%) | 82195 (1·3%) | 29064 (0·8%) | 907323 (1·5%) |
| **Atrial fibrillation** | 1300604 (2·5%) | 173928 (2·7%) | 36506 (1·0%) | 1511038 (2·4%) |
| **Acute myocardial infraction** | 872067 (1·7%) | 99550 (1·5%) | 20305 (0·5%) | 991922 (1·6%) |
| **Chronic kidney disease** | 2147541 (4·2%) | 245624 (3·7%) | 59129 (1·6%) | 2452294 (4·0%) |
| **COPD** | 1123202 (2·2%) | 104455 (1·6%) | 24733 (0·7%) | 1252390 (2·0%) |
| **Heart failure** | 701978 (1·4%) | 88202 (1·3%) | 16738 (0·4%) | 806918 (1·3%) |
| **Pulmonary embolism** | 5257 (0·0%) | 529 (0·0%) | 129 (0·0%) | 5915 (0·0%) |
| **Cancer** | 6203838 (12·0%) | 604350 (9·2%) | 208500 (5·5%) | 7016688 (11·4%) |
| **Dementia** | 501322 (1·0%) | 76545 (1·2%) | 16203 (0·4%) | 594070 (1·0%) |
| **Diabetes** | 3703702 (7·2%) | 321356 (4·9%) | 106695 (2·8%) | 4131753 (6·7%) |
| **Hypertension** | 7860172 (15·3%) | 809692 (12·3%) | 252239 (6·7%) | 8922103 (14·4%) |
| **Liver disease** | 174646 (0·3%) | 17650 (0·3%) | 4649 (0·1%) | 196945 (0·3%) |
| **Obesity** | 2611818 (5·1%) | 241761 (3·7%) | 82667 (2·2%) | 2936246 (4·8%) |
| **Stroke** | 1048990 (2·0%) | 131630 (2·0%) | 27982 (0·7%) | 1208602 (2·0%) |

*\*Excluding individuals who refused to state their ethnicity (NHS ethnicity code was 'Z').*
*\*\*Group composed by individuals who refused to state their ethnicity (NHS ethnicity code was 'Z') and those whose ethnicity was not recorded.*
*\*\*\*Geographic regions reported in the table belongs to the nine official regions of England. Abbreviations: COPD, chronic obstructive pulmonary disease; IMD, index of multiple deprivation; NHS, National Health Service in the UK; SD, standard deviation.*

### Multiple records

About 1·4% of individuals with an original NHS ethnicity code record and 16·0% of individuals with a converted SNOMED concept record had multiple different ethnicity codes (Table 3). Excluding the *Not stated* (Z) code reduced inconsistences. In contrast, 38·0% of individuals in GDPPR with at least one ethnicity record in HES (n=46,804,958) had multiple inconsistent records, dropping to 19·0% when the *Not stated* (Z) code was excluded.

**Table 3.** Discrepancies in multiple ethnicity records across GDPPR and HES tables using individuals identified in GDPPR.

| Ethnicity data source and code type used | Individuals with recorded ethnicity* (n) | Individuals with inconsistent multiple records (n, %) |  | Individuals with inconsistent multiple records, excluding "Z - Not stated" code (n, %) |  |
| --- | --- | --- | --- | --- | --- |
| All GDPPR Codes | 53,266,674 | 9,379,041 | 17.0% | 6,453,095 | 12.0% |
| GDPPR NHS ethnicity Code | 29,721,460 | 426,194 | 1.4% | 353,828 | 1.2% |
| GDPPR SNOMED concept | 51,135,903 | 8,163,018 | 16.0% | 5,260,197 | 10.3% |
| Linked-HES NHS ethnicity Code | 46,804,958 | 17,704,692 | 38.0% | 8,658,447 | 19.0% |

*\*Including NHS ethnicity code 'Z – not stated' and the SNOMED concepts mapped to it. Abbreviations: GDPPR, General Practice Extraction Service (GPES) Data for Pandemic Planning and Research; NHS, National Health Service in the UK; HES, hospital episode statistics; SNOMED, SNOMED-CT records containing ethnicity concepts.*

The most common ethnicity codes when multiple codes were recorded in GDPPR were *British* (A), *Any other White background* (C), *Not stated (Z), Any other ethnic group* (S), and *Any other Asian background* (L) (Figure 3).

**Figure 3.**
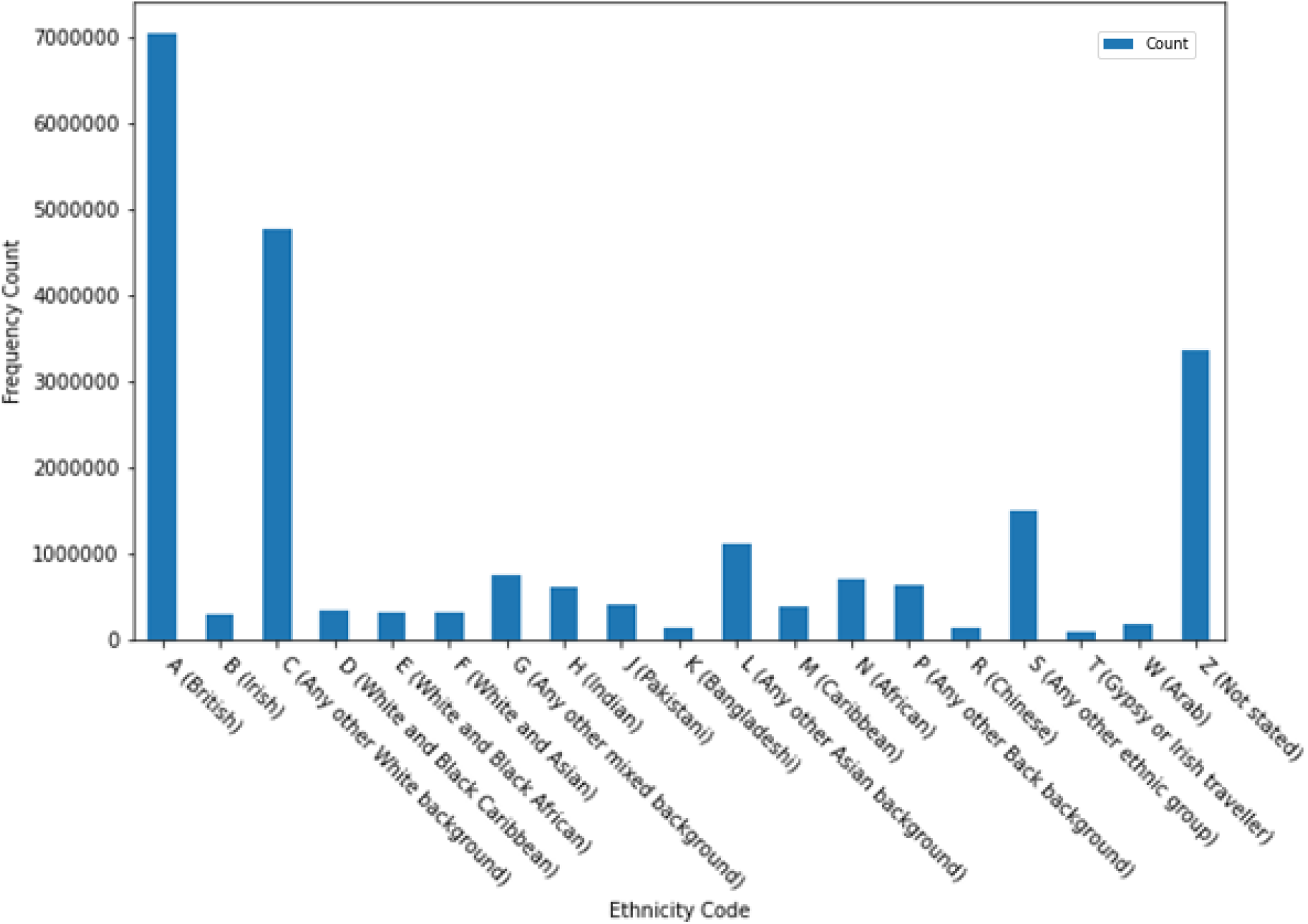
Ethnicity codes that were most frequently found in records with more than one reported code.

The most common ethnicity code pairings were *British* (A) – *Any other White background* (C), *British* (A) – *Not stated* (Z), and *Any other White background* (C) – *Any other ethnic group* (S). When White ethnicity codes (A, B, and C) were excluded, the most common pairs of minority ethnic codes were *Any other Asian background* (L) – *Any other ethnic group* (S), *African* (N) – *Any other Black background* (P), and Indian (H) *– Any other Asian background* (L) (Figure 4).

**Figure 4.**
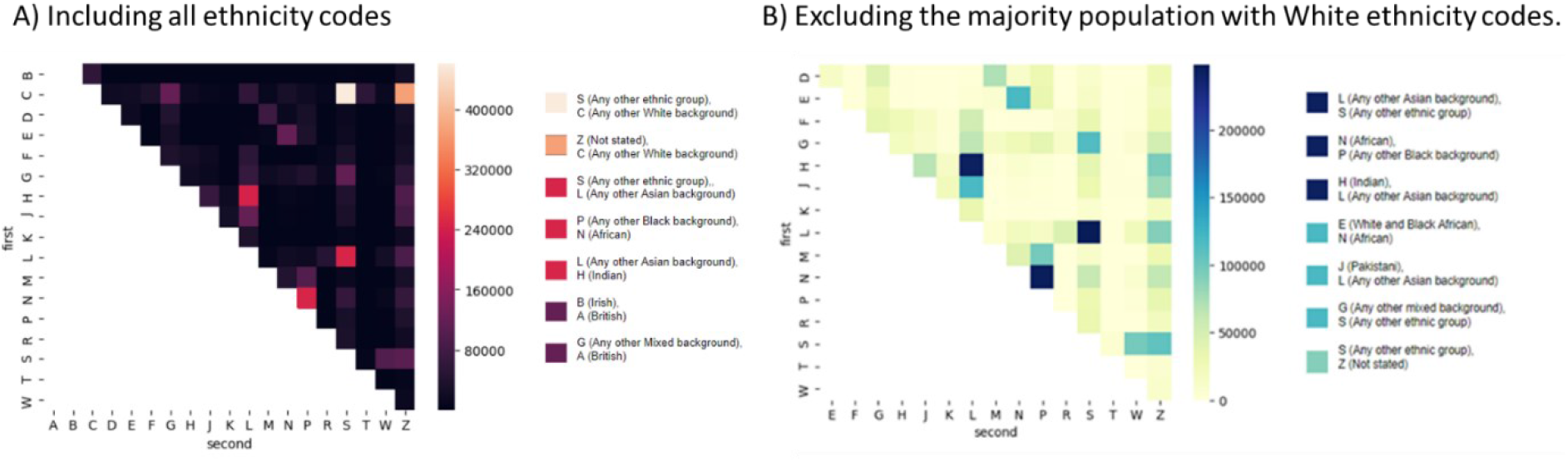
Frequency of pairwise occurrence when A) including all ethnicity codes and B) excluding the majority population with White ethnicity codes (A, B or C). NHS ethnicity codes: A, British; B, Irish; C, Any other White background, D, White and Black Caribbean; E, White and Black African; F, White and Asian; G, Any other mixed background; H, Indian; J, Pakistani; K, Bangladeshi; L, Any other Asian background; M, Caribbean; N, African; P, Any other Black background; R, Chinese; S, Any other ethnic group; T, Traveller; W, Arab; Z, Not stated. Abbreviations: NHS, National Health Service in the UK.

### Granularity of ethnicity data

Figure 5 maps the distinct levels of ethnicity concepts from the different data sources to one another. SNOMED currently gives the most granular ethnicity records (Supplemental file “GDPPR Tree diagram including all categories.html”), with 489 SNOMED concepts representing ethnicity within the NHS Digital TRE (*Appendix SNOMED mapping*). However, only 255 (52·1%) of these codes were used at least once in the extracted individuals’ records. The remaining 234 (47·8%) codes were not assigned to any individual (Figure 6). Figure 7 shows the five most frequently used SNOMED concepts mapped to each NHS ethnicity code. *Appendix SNOMED GDPPR counts* displays the number of individuals per SNOMED concept in a table format.

**Figure 5.**
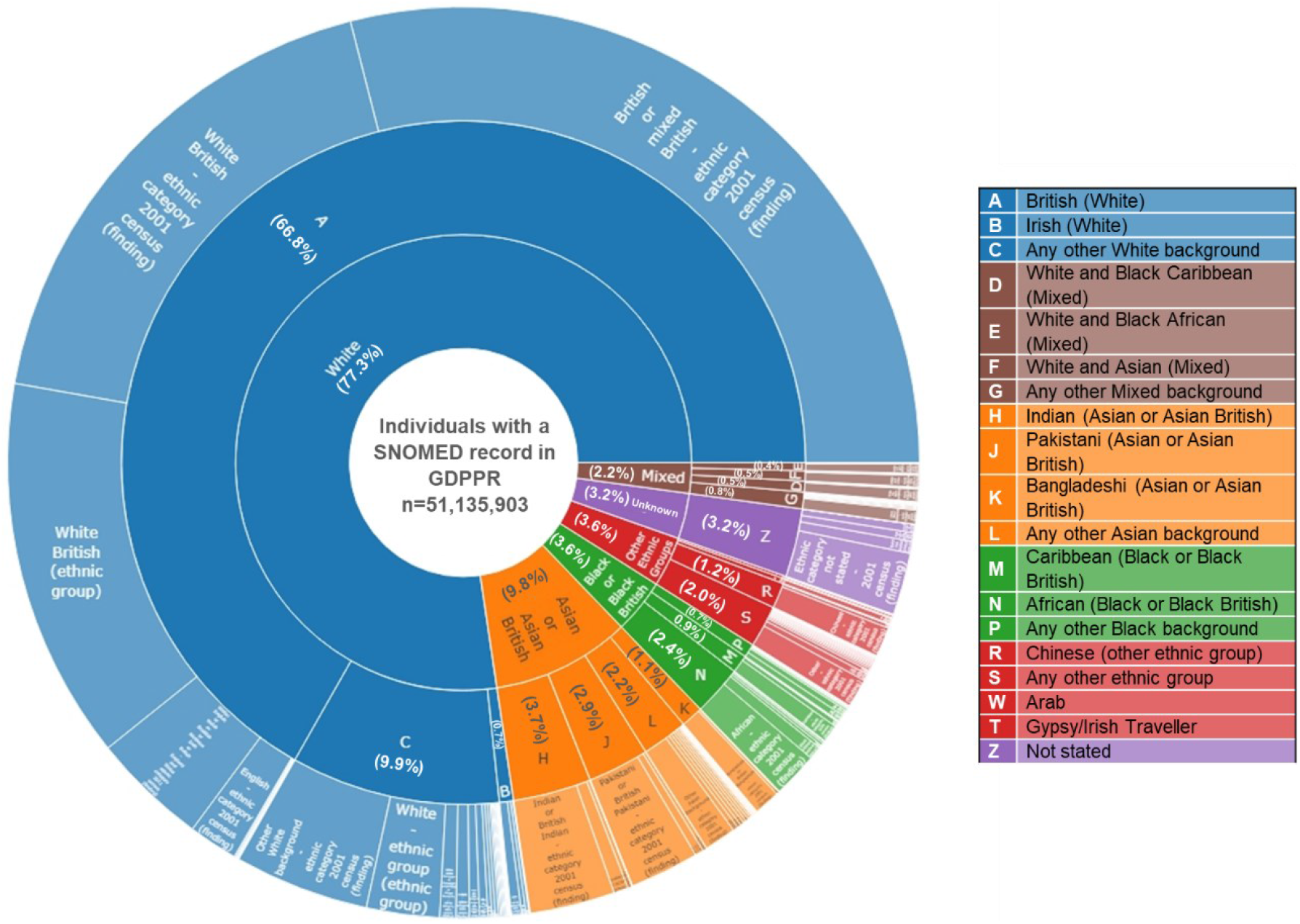
Representation of the hierarchical structure from “high-level ethnic groups” to NHS ethnicity Codes and the observed 255 SNOMED concepts. The “observed SNOMED concepts” refers to the 255 of 489 concepts available in the TRE that were used at least once in the GDPPR dataset (n = 51,135,903). Branches from the same high-level ethnic groups are coloured using the same colour or shade. Abbreviations: High-level ethnic groups, general ethnicity classification groups from the Office for National Statistics commonly used in research; NHS, National Health Service in the UK; SNOMED, SNOMED-CT records containing ethnicity concepts; TRE, NHS Digital Trusted Research Environment.

**Figure 6.**
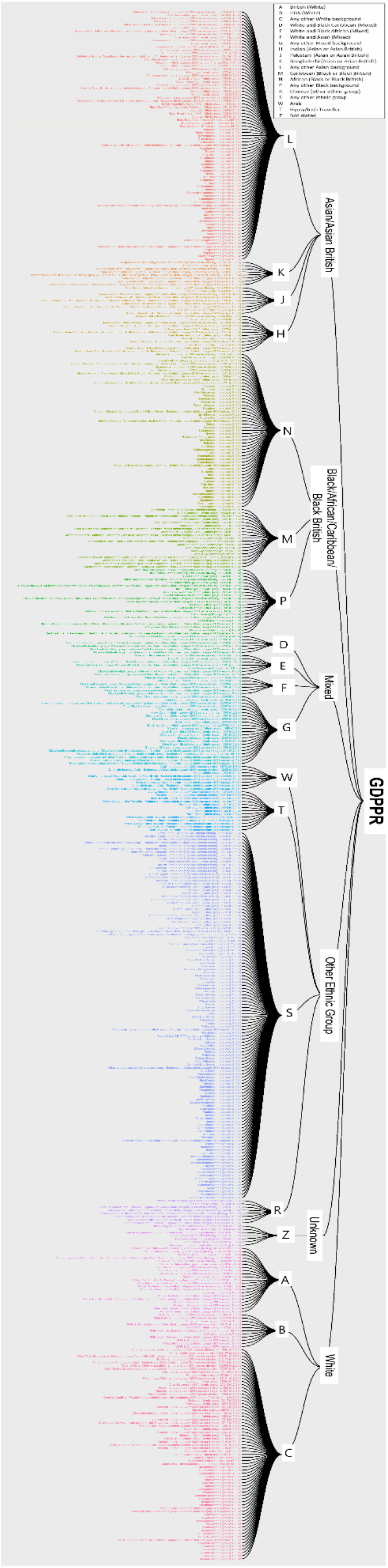
Tree diagram representing the mapping between “high-level ethnic groups”, NHS ethnicity Codes, and the observed 255 SNOMED concepts. The “observed SNOMED concepts” refers to the 255 of 489 concepts available in the TRE that were used at least once in the GDPPR dataset, including “non-stated” codes (n = 51,135,903). SNOMED concepts belonging to the same NHS ethnicity code are represented using the same colour. Abbreviations: High-level ethnic groups, general ethnicity classification groups from the Office for National Statistics commonly used in research; NHS, National Health Service in the UK; SNOMED, SNOMED-CT records containing ethnicity concepts; TRE, NHS Digital Trusted Research Environment.

**Figure 7.**
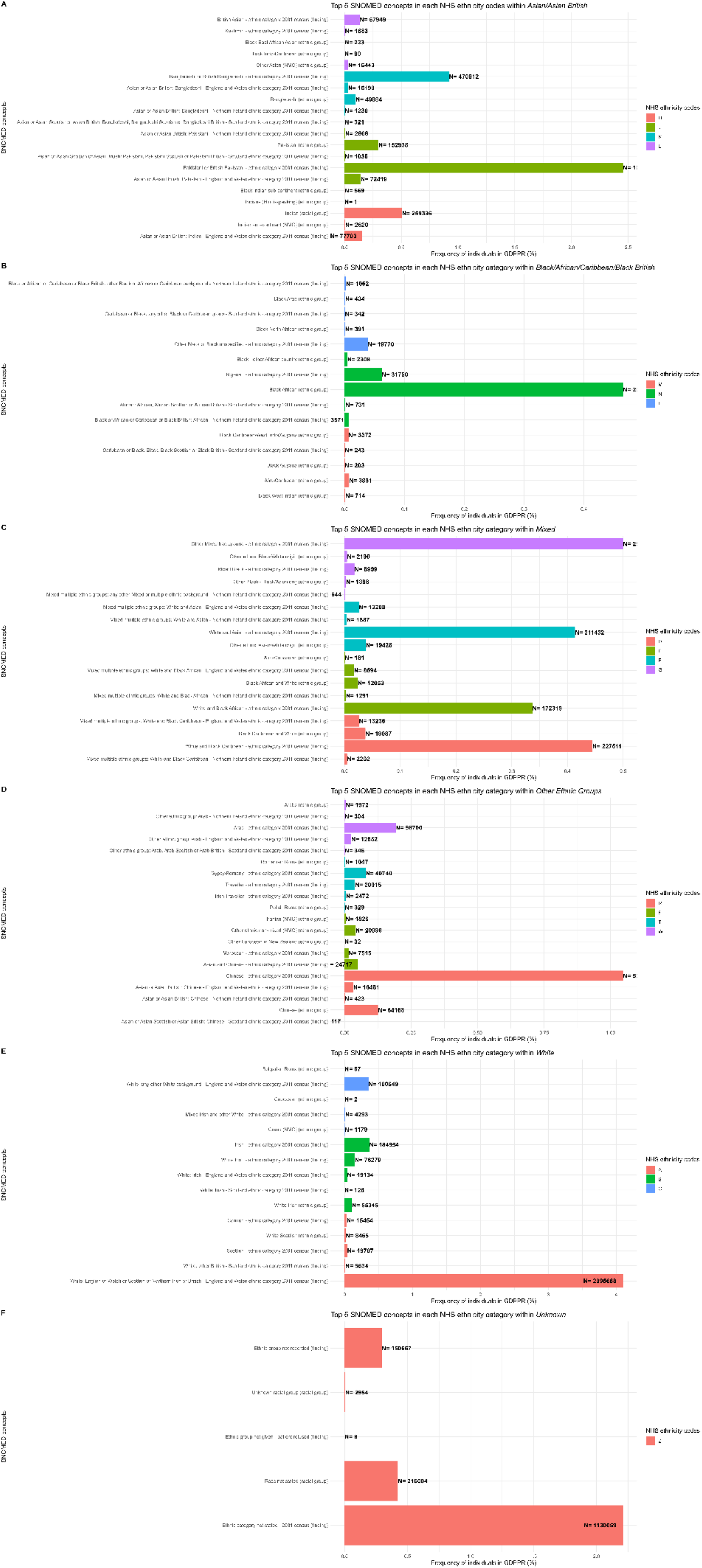
Top five SNOMED concepts most frequently used in each NHS ethnicity Code level, arranged by “high-level ethnic groups”: A) Asian/Asian British, B) Black/African/Caribbean/Black British, C) Mixed, D) Other Ethnic Groups, E) White and F) Unknown. NHS ethnicity codes: A, British; B, Irish; C, Any other White background, D, White and Black Caribbean; E, White and Black African; F, White and Asian; G, Any other mixed background; H, Indian; J, Pakistani; K, Bangladeshi; L, Any other Asian background; M, Caribbean; N, African; P, Any other Black background; R, Chinese; S, Any other ethnic group; T, Traveller; W, Arab; Z, Not stated. Abbreviations: High-level ethnic groups, general ethnicity classification groups from the Office for National Statistics commonly used in research; NHS, National Health Service in the UK; SNOMED, SNOMED-CT records containing ethnicity concepts.

### Diversity in SNOMED concepts

SNOMED concepts were substantially diverse. Of the 255 codes in use, 162 (63·5%) contained an ethnicity/race concept, 5 (2·0%) included a religion, 187 (73·3%) referenced a geographical region, and 60 (23·5%) referenced a language. Full list is available at Appendix SNOMED diversity.

### Potential discrepancies and misclassifications

Some inconsistency was found in the aggregation of NHS ethnicity categories into the high-level ethnic groups (Figure 6). According to the 2011 and 2021 England and Wales census classifications, *Gypsy/Irish Traveller* (T) falls within the higher-level category *White*, and *Chinese* (R) within *Asian*. In contrast, the NHS ethnicity classification included *Gypsy/Irish Traveller* (T) and *Chinese* (R) within the higher-level category *Other Ethnic Groups*, following the ONS Census 2001 classification (see classification algorithm used in the CVD-COVID-UK/Covid impact consortium in supplemental data file).

Further discrepancies were found in the grouping of SNOMED concepts into NHS ethnicity categories. A mapping algorithm could not be traced. Given the lack of documentation on the mapping of SNOMED concepts to NHS ethnicity code, several potential discrepancies were observed which should be carefully (re)considered by researchers in future (Figure 8): for instance, concepts including a variant of “Black East African Asian/Indo-Caribbean” were assigned to *Any other Asian background* (L). However, there is no clarification as to whether these concepts are mapped more accurately there than other groups as Other ethnic group or Black/African/Caribbean/Black British. Likewise, variants of “Black West Indian” were mapped to *Caribbean* (M), although a proportion of these individuals may have Asian legacy. Arguably, the three ‘Mixed’ concepts within *Any other background* (G) may be better grouped in more specific categories, such as “Black -other, mixed” within *Black background* (P). Several concepts contained by *Any other ethnic group* (S) could also be placed in more specific categories. For example, the 2001 census category “Asian and Chinese” is linked to *Any other ethnic group* (S) instead of *Chinese* (R) or *Any other Asian background* (L). Similarly, some SNOMED concepts include the concept “Roma”. As the ONS 2021 census included the new category *White: Roma*, the mapping could be updated to reflect this change.

**Figure 8.**
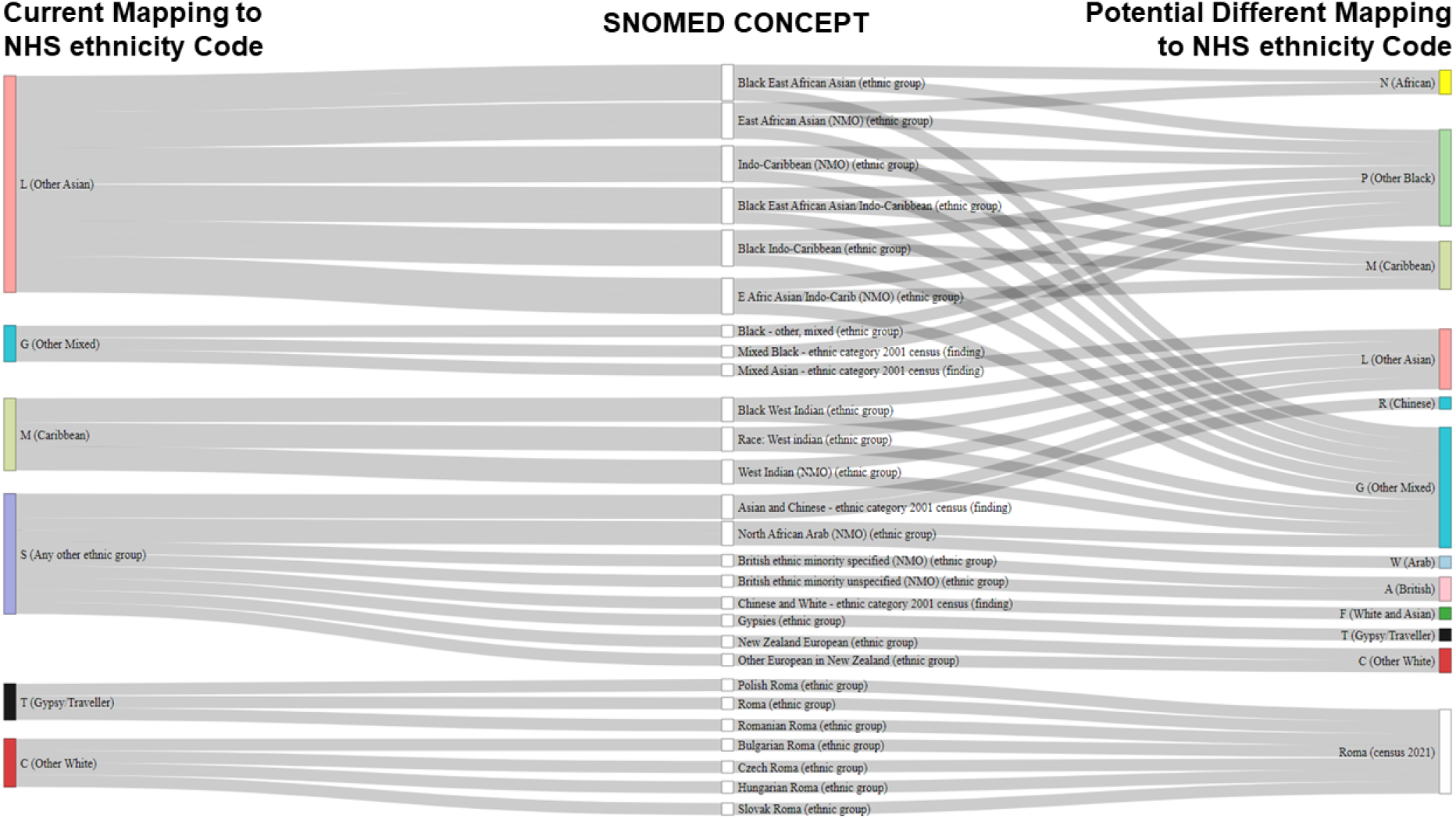
Sankey plot showing potential discrepancies between SNOMED concepts and NHS ethnicity codes mapping. Abbreviations: NHS, National Health Service in the UK; SNOMED, SNOMED-CT records containing ethnicity concepts.

## Discussion

Biased ethnicity knowledge can lead to biased healthcare decision-making and to patients receiving inappropriate or no care, thereby negatively affecting their outcomes. It can also increase costs to the care system and affect public trust. Identification of ethnicity is an essential first step to understanding inequities between ethnicities. Despite its complexity, researchers should aim to include ethnicity in their analyses. The results presented here can be used to further the use of ethnicity in future research.

### Completeness of ethnicity records

We found that over 83·3% of individuals in England primary care system had at least one ethnicity recorded; increasing to 93·9% when linked to hospitalisation records. This result is consistent with prior findings^15^ and highlights the usefulness of linking data across primary and secondary care to maximise ethnicity data completeness. Individuals with missing ethnicity were younger, more likely to be male and living in the southern regions of England, and had fewer comorbidities than individuals with recorded ethnicity. Similar results have been reported in other UK data sources. For instance, Mathur et al. observed higher rates of ethnicity records for individuals aged 40 to 79 years in the Clinical Practice Research Datalink (CPRD) and HES data sources than older or younger individuals.^8^ Petersen at al. found that, among people aged 18-65 years, men were less likely to have health indicators recorded than women in the Health Improvement Network (THIN).^22^

### Granularity of ethnicity data

Most studies collapse the available ethnicity concepts into five (e.g., Asian, Black/African/Caribbean, White, Mixed, Other Ethnic Groups) or six (the previously mentioned plus Unknown ethnicity) categories.^11^ However, some studies have accounted for greater distinctions by exploring ethnic minorities such as Bangladeshi in the UK and Hispanic/Latinos in the US.^23^ This work describes for the first time more than 250 patient-identified ethnicity sub-groups in England.

Ethnicity data in healthcare and national statistics are captured for different purposes, partly explaining some of the differences among NHS and census ethnicity categories. For instance GDPPR is directly intended for patient care; HES has a more administrative nature linked to payments; while the ONS collects information from the UK population, including ethnicity, in a census held approximately every 10 years, most recently in 2021^7,20^. To allow for the emergence of new ethnic groups, the census questionnaire allows free-text answers.^20^ After pooling all information, the ONS reports the groups that, in their understanding, best represent the existing diversity in the UK. Updated ethnicity groups are then shared with the NHS, which uses this information to update the ethnicity categories used in their data sources.

The 2011 Census published by the ONS was the gold standard for ethnicity recording in England and Wales until the recent publication of the 2021 Census.^24^ However, not all NHS sources base their categories on the same census. For example, HES uses 2001 Census categories, whereas GDPPR uses 2011 census categories. This discrepancy creates uncertainties and data mismatching between different datasets. For example, HES does not include the ethnicity categories *Arab* (W) and *Gypsy/Irish Traveller* (T).^25^ This highlights once more the importance of linking data across primary and secondary care, in this case, to maximise ethnicity data granularity.

Despite differences, we can compare the prevalence of SNOMED concepts used in GDPPR to the latest population estimates in England and Wales (2019).^26^ Fewer individuals self-identified as White British (66·8% in GDPPR vs· 78·4% in 2019 estimates), Gypsy/Traveller (0·1% vs· 0·03%), or Arab (0·2% vs 0·4%). Higher proportion of individuals self-identified as Chinese (1·2% vs 0·6%), Indian (3·7% vs· 2·8%), Pakistan (2·9% vs 2·3%), or Any other mixed background (0·8% vs 0·5%). And similar percentage self-identified as Bangladeshi (1·1% vs 1·0%), Caribbean (0·9% vs 1·0%), African (2·4% vs 2·3%), White and Asian (0·5% both), White and Black African (0·4% vs 0·3%), or White and Black Caribbean (0·5% both). The 2019 census did not include people who had died or individuals with an unknown ethnicity, which might account for these differences.

### Multiple records and potential discrepancies

Our analysis of multiple ethnicity records within GDPPR and HES sources showed similar discrepancy rates when the code for “do not know/refusal” (i.e., *Not stated* (Z)) was excluded. However, GDPPR ethnicity data should be prioritised to ensure inclusion of *Arab* (W) and *Gypsy/Irish Traveller* (T) ethnicities. Within patient records, the most frequently coexisting codes would be placed in the same higher-order group. For example, codes for *African* (N) and *Any other Black background* (P) often appear for the same individual and would both be grouped within the high-level category Black/African/Caribbean/Black British. The use of higher-level groupings can therefore resolve some conflicting cases by reducing granularity. However, it cannot resolve conflicts where different Mixed categories coexist in the same record, such as *Any Other Mixed Background* (G) occurring alongside *British* (A), *Any Other White Background* (C), or *African* (N). Higher-level Mixed groupings may therefore include more ambiguous ethnicity concepts. The *British* (A) code had frequent conflicting pairings with *Indian* (H), *Any other Asian background* (L), and *Caribbean* (M), suggesting inconsistencies in individuals’ perceptions of their nationality and ethnicity when self-reporting ethnicity.

The grouping algorithm used can also be a source of inconsistencies. For instance, including *Chinese* (R) and *Gypsy/Irish Traveller* (T) within Other Ethnic Groups instead of the established high-level ethnic groups might be preferred for certain studies, but should not be by default.

Uncertainty regarding mapping of international SNOMED-CT ethnicity concepts to NHS ethnicity codes highlighted the need for better documentation of underlying processes. Several potential discrepancies were observed e.g., “Asians plus African/Black/Caribbean” minorities (e.g., “Black East African Asian/Indo-Caribbean (ethnic group)”) were all linked with the NHS ethnicity code *Any other Asian background* (L). Historically, the “Asians plus African/Black/Caribbean” concepts often refer to individuals from the Indian subcontinent who migrated to Africa, and then from Africa to the UK.^27^ However, SNOMED concepts available in GDPPR data account for different, more granular ethnic groups than NHS ethnicity codes, enabling greater diversity in ethnicity groups to be represented. The descriptions of the observed SNOMED concepts included ethnicity, race, religion, and geographic location, among others. However, many concepts require some aggregation due to their limited use within very large datasets, such as the one explored here. Most research based on NHS data uses wider categories, rather than the highly specific concepts captured by the SNOMED concepts. The large variety and complexity of ethnicity codes can make collapsing and comparing codes difficult, regardless of whether NHS ethnicity codes or high-level ethnic groups are used. Although using these more general groupings allows researchers to achieve a minimum sample size while protecting individual identities, the cost is the uncertainty of how accurate these bigger groups are.

#### Strengths and limitations of this study

GDPPR is a collection of de-identified person-level primary care data (linked to secondary and tertiary care) for one of the world’s largest population-wide electronic health records database and housed within a trusted research environment, NHS Digital for England. This study provides a greater and deeper understanding of the use of ethnicity in this dataset. The observed findings are highly representative of the England population. However, there are some potential limitations.

There is no perfect solution for conflicting codes in the same individual, especially for codes that cannot be reconciled (e.g., White, Black). Of the different available approaches, we used the most recent SNOMED concepts in an individual’s record when exploring granularity in GDPPR. This approach may have affected the prevalence of very small minority groups, including the 234 codes that were not linked to any user. However, using the most recent codes allowed us to include any new ethnicity definitions within SNOMED.

The Master Person Service algorithm increases the quality of the data by matching and linking person-records within and across the different NHS sources.^28^ However, presence of individuals duplicated is still feasible. Further studies are required to assess its accuracy in GDPPR records.

Whilst improvements to data collection at source would be welcome, much more can be done with currently available ethnicity data than is typically seen in the literature. This is important to be done now more than ever as routinely collected ethnicity data are increasingly used in the era of real-world analytics and large-scale trials. This study demonstrates the importance of linking data across primary and secondary care to maximise both ascertainment/completeness and granularity of ethnicity data, and application of better ethnicity coding in big health data.

## Conclusion

To our knowledge this work is the first study on over 250 patient-identified ethnicity sub-groups among over 61 million individuals in England. Ethnic diversity is better captured in SNOMED concepts than other existing classifications. Study-specific sample size considerations may require combining smaller ethnicity categories into larger groups. Improvements required in ethnicity mapping between classifications were identified.

Using the results of this study, researchers can use more granular ethnicity than the typical approaches which aggregate ethnicity into a limited number of categories, failing to reflect the diversity of underlying populations. Accurate ethnicity data will lead to a better understanding of individual diversity, which will help to address disparities and influence policy recommendations that can translate into better, fairer health for all.

## Data Availability

The data used in this study are available in NHS Digital's TRE for England but, as restrictions apply, they are not publicly available (https://digital.nhs.uk/coronavirus/coronavirus-data-services-updates/trusted-research-environment-service-for-england). The CVD-COVID-UK/COVID-IMPACT programme led by the BHF Data Science Centre (https://www.hdruk.ac.uk/helping-with-health-data/bhf-data-science-centre/) received approval to access data in NHS Digital's TRE for England from the Independent Group Advising on the Release of Data (IGARD) (https://digital.nhs.uk/about-nhs-digital/corporate-information-and-documents/independent-group-advising-on-the-release-of-data) via an application made in the Data Access Request Service (DARS) Online system (ref. DARS-NIC-381078-Y9C5K) (https://digital.nhs.uk/services/data-access-request-service-dars/dars-products-and-services). The CVD-COVID-UK/COVID-IMPACT Approvals & Oversight Board (https://www.hdruk.ac.uk/projects/cvd-covid-uk-project/) subsequently granted approval to this project to access the data within NHS Digital's TRE for England. The de-identified data used in this study were made available to accredited researchers only. Those wishing to gain access to the data should contact in the first instance.

https://github.com/BHFDSC/CCU037_01

## Acknowledgments

The British Heart Foundation Data Science Centre (grant No SP/19/3/34678, awarded to Health Data Research (HDR) UK), funded co-development (with NHS Digital) of the trusted research environment, provision of linked datasets, data access, user software licences, computational usage, and data management and wrangling support, with additional contributions from the HDR UK Data and Connectivity component of the UK Government Chief Scientific Adviser’s National Core Studies programme to coordinate national COVID-19 priority research. Consortium partner organisations funded the time of contributing data analysts, biostatisticians, epidemiologists, and clinicians. The authors acknowledge English language editing by Dr Jennifer A de Beyer, Centre for Statistics in Medicine, University of Oxford.

This work was carried out with the support of the BHF Data Science Centre led by HDR UK (BHF Grant no. SP/19/3/34678). This study made use of de-identified data held in NHS Digital’s TRE for England and made available via the BHF Data Science Centre’s CVD-COVID-UK/COVID-IMPACT consortium. This work uses data provided by patients and collected by the NHS as part of their care and support. We would like to acknowledge all data providers who make health relevant data available for research.

This research is part of the Data and Connectivity National Core Study, led by Health Data Research UK in partnership with the Office for National Statistics and funded by UK Research and Innovation (grant ref MC_PC_20058). This work was also supported by The Alan Turing Institute via ‘Towards Turing 2.0’ EPSRC Grant Funding.

## Authors’ roles

Conceptualisation: SK, DPA, AD, GC. Data curation: MPM, FA. Formal analysis: MPM, FA, TB. Funding acquisition: SK. Data interpretation: MPM, FA, SK. Writing original draft: MPM, FA, SK. Writing review and editing: all authors. Approving final version of manuscript: all authors. SK and MPM takes responsibility for the integrity of the data analysis. CS is the Director of the BHF Data Science Centre and coordinated approvals for and access to data within NHS Digital’s TRE for England for CVD-COVID-UK/COVID-IMPACT.

## Ethical approval

The North East - Newcastle and North Tyneside 2 research ethics committee provided ethical approval for the CVD-COVID-UK/COVID-IMPACT research programme (REC no: 20/NE/0161) to access, within secure trusted research environments, unconsented, whole-population, de-identified data from electronic health records collected as part of patients’ routine healthcare.

Our project (proposal CCU037, short title: *Minimising bias in ethnicity data*) agreed the objectives of the consortium’s ethical and regulatory approvals and was authorised by the BHF Data Science Centre’s Approvals and Oversight Board. Approved researchers (MPM, AD, SK) conducted the analyses within the NHS Digital TRE via secure remote access. Ensuring the anonymity of individuals, only summarised-aggregated results that were manually reviewed by the NHS Digital ‘safe outputs’ escrow service were exported from the TRE environment.

## Data sharing

The data used in this study are available in NHS Digital’s TRE for England but, as restrictions apply, they are not publicly available (<https://digital.nhs.uk/coronavirus/coronavirus-data-services-updates/trusted-research-environment-service-for-england>). The CVD-COVID-UK/COVID-IMPACT programme led by the BHF Data Science Centre (https://www.hdruk.ac.uk/helping-with-health-data/bhf-data-science-centre/) received approval to access data in NHS Digital’s TRE for England from the Independent Group Advising on the Release of Data (IGARD) (<https://digital.nhs.uk/about-nhs-digital/corporate-information-and-documents/independent-group-advising-on-the-release-of-data>) via an application made in the Data Access Request Service (DARS) Online system (ref. DARS-NIC-381078-Y9C5K) (https://digital.nhs.uk/services/data-access-request-service-dars/dars-products-and-services). The CVD-COVID-UK/COVID-IMPACT Approvals & Oversight Board (https://www.hdruk.ac.uk/projects/cvd-covid-uk-project/) subsequently granted approval to this project to access the data within NHS Digital’s TRE for England. The de-identified data used in this study were made available to accredited researchers only. Those wishing to gain access to the data should contact in the first instance.

