## Supplementary material for "Digital ethnicity data in population-wide electronic health records in England: a description of completeness, coverage, and granularity of diversity": Supplemental data.docx

**Methods - further details:**

### Data sources and linkages

NHS Digital maintains a TRE for secure access to anonymised patient-level electronic health records for England with linkages to primary-, secondary-, and tertiary-care data sources for research purposes.^1,2^ The NHS Digital’s Master Person Service facilitates the linkage between the TRE data sources though the NHS number (a unique 10 digit healthcare identifier), date of birth, and sex.^1,3^

This study focused on the General Practice Extraction Service (GPES) Data for Pandemic Planning and Research (GDPPR) data sources, a primary-care dataset for England that collects information from all individuals who are currently registered with a general practitioner (GP) practice and any individual who died on or after 1^st^ November 2019. Individuals registered with a GP in England who died before November 2019 are not included in the GDPPR. Data include diagnoses, prescriptions, treatments, outcomes, vaccinations, and immunisations.^4,5^ GDPPR covers 98% of English GP practices across all relevant GP computer system suppliers (TPP, EMIS, Cegedim (formerly called Vision or In Practice Systems), and Microtest).^1^

### Other ethnicity classifications - High-level ethnic groups

**Algorithm to collapse NHS ethnicity codes into the six high-level ethnic categories*:**

CASE WHEN ETHNICITY_CODE IN ('1','2','3','N','M','P') THEN "Black or Black British"

           WHEN ETHNICITY_CODE IN ('0','A','B','C') THEN "White"

           WHEN ETHNICITY_CODE IN ('4','5','6','L','K','J','H') THEN "Asian or Asian British"

           WHEN ETHNICITY_CODE IN ('7','8','W','T','S','R') THEN "Other Ethnic Groups"

           WHEN ETHNICITY_CODE IN ('D','E','F','G') THEN "Mixed"

           WHEN ETHNICITY_CODE IN ('9','Z','X') THEN "Unknown"

           ELSE 'Unknown' END as ETHNIC_GROUP

*This is the regularly used algorithm within the CVD-COVID-UK/Covid impact consortium. Codes for *Gypsy/Irish Traveller* ('T') and Chinese ('R') should be placed within "White" and "Asian or Asian British", respectively, according to the 2011 and 2021 ONS Census.

### Covariates - additional characteristics

Additional characteristics of individuals extracted from GDPPR data included: age at date of death or age on 23^rd^ April 2022, sex, most recent record of residence (i.e., geographical region) in England, body mass index (BMI), index of multiple deprivation (IMD), current smoking status, current alcohol use status, and the presence of any clinical record of atrial fibrillation, acute myocardial infarction, chronic kidney disease, chronic obstructive pulmonary disease (COPD), heart failure, pulmonary embolism, cancer, dementia, diabetes, hypertension, liver disease, obesity, or stroke diagnosis. Geographical region is reported using England’s nine official regions: London, North East, North West, Yorkshire, East Midlands, West Midlands, South East, East, and South West; which were mapped from the Lower Layer Super Output Areas (LSOA).
