## Supplementary material for "Digital ethnicity data in population-wide electronic health records in England: a description of completeness, coverage, and granularity of diversity": Supplemental figures - Ethnicity GDPPR.docx

### Supplemental Figure 1. Decision tree of preferred source of ethnicity.

Solid arrows mark the preferred option whilst dashed arrows indicate the alternative route.

Abbreviations: GDPPR, General Practice Extraction Service (GPES) Data for Pandemic Planning and Research; HES, hospital episode statistics; SNOMED, SNOMED-CT records containing ethnicity codes.

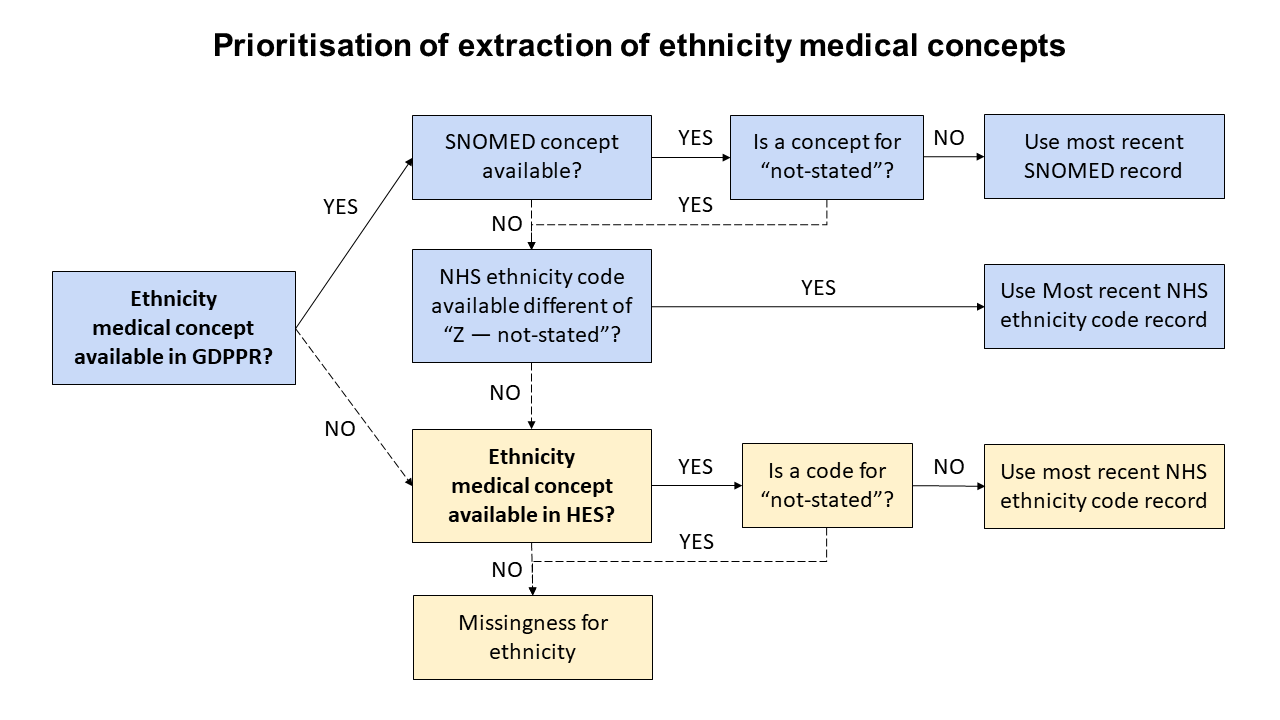

### Supplemental Figure 2. England regions with percentage of individuals with no ethnicity records.

In GDPPR, 14.7 % did not have a post code record and could not be allocated in the map.
The shown values are the frequency of “individuals with no ethnicity divided by the total individuals” in each of the England regions.

Abbreviations: GDPPR, General Practice Extraction Service (GPES) Data for Pandemic Planning and Research.

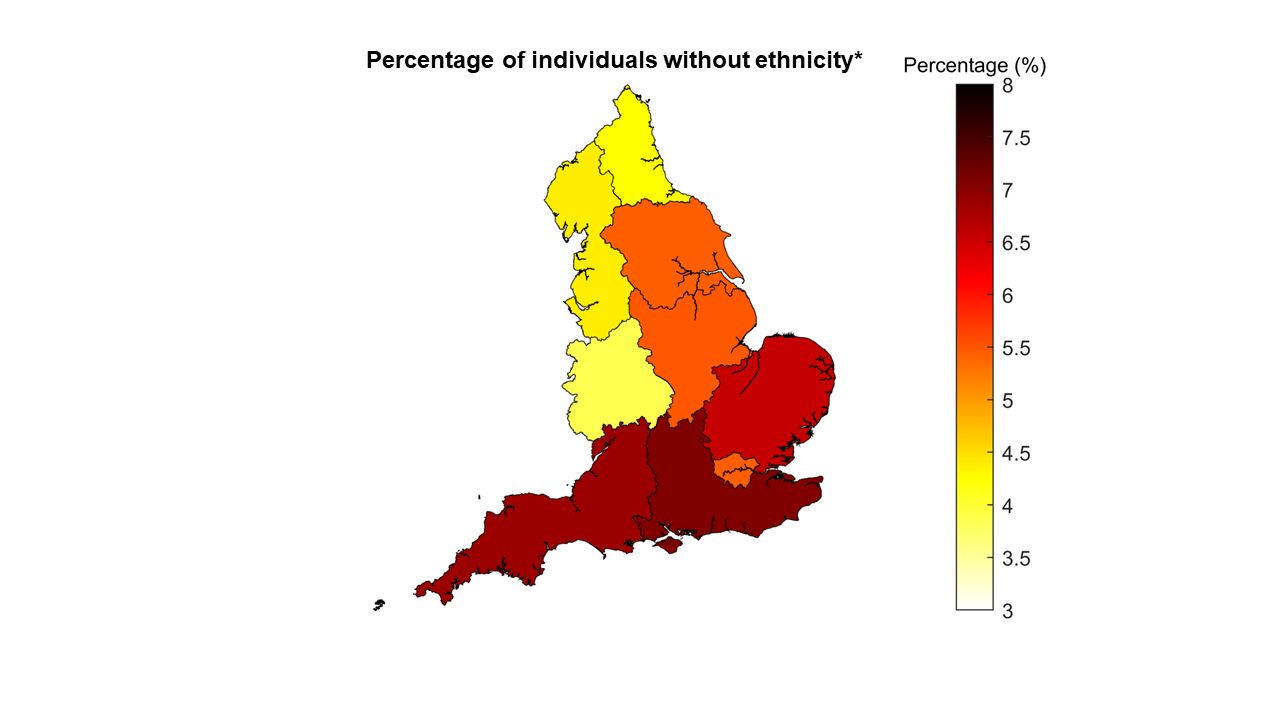
